# The concentration of tuberculosis within Paraguay’s incarcerated and Indigenous populations, 2018-2022

**DOI:** 10.1101/2024.05.31.24308287

**Authors:** Angélica Medina, Jacob Sussman, Natalia Sosa, Melissa Valdez, Jason R. Andrews, Julio Croda, Gladys Estigarribia Sanabria, Guillermo Sequera, Sarita Aguirre, Katharine S. Walter

## Abstract

While incidence of tuberculosis (TB) has decreased globally, in Paraguay, considered a medium-incidence country by the WHO, TB incidence has increased slightly from 42 per 100,000 in 2010 to 46 per 100,000 in 2022. We conducted a retrospective study of TB cases notified to the Paraguay National Program for Tuberculosis Control (NPTC) from 2018 to 2022 and quantified trends in specific populations identified as vulnerable. Of the 13,725 TB cases notified in Paraguay from 2018 to 2022, 2,331 (17%) occurred among incarcerated individuals and 1,743 (12.7%) occurred among self-identified Indigenous individuals. In 2022, the relative risk of TB was 87 and 6.4 among the incarcerated and Indigenous populations, compared with the non-incarcerated and non-Indigenous populations respectively. We found significant heterogeneity in TB incidence across Paraguay’s 17 departments. Our findings highlight the urgency of expanding access to TB diagnosis, treatment, and prevention in populations at heightened risk of TB in Paraguay.

## INTRODUCTION

Tuberculosis (TB) remains the leading cause of death from an infectious disease globally (1). Despite being both preventable and curable, 1.3 million people died from TB in 2022 (1).

While over the past decade global incidence rates of TB have decreased, between 2015 and 2022 there was a 13% increase in TB incidence in the Americas (2,3). In Paraguay, a medium- incidence country, the TB incidence increased from 42 (37-48) per 100,000 in 2010 (2) to 46 (39-54) per 100,000 in 2022 (4,5). Recently observed increases in TB notifications may partly reflect the impact of the COVID-19 pandemic on TB case detection and treatment. Yet what explains recent increases in the country’s TB notifications and whether specific populations remain at heightened risk of TB is not fully understood.

Globally, TB is a disease of poverty and is concentrated among populations marginalized from access to healthcare and other social services. For example, incarcerated populations have an incidence nine times that of the non-incarcerated population, and the relative risk of TB among incarcerated populations is many times higher in some countries (6). This reflects extreme overcrowding, lack of or limited access to healthcare, poor access to nutrition, and other characteristics of carceral environments. In the Americas, Indigenous populations are also at heightened risk for TB, reflecting historic and ongoing displacement and exclusion from access to healthcare and other services (7,8). TB incidence among Indigenous peoples in South America reached epidemic levels following European colonization (7), and has been amplified by social marginalization, food insecurity, and a lack of access to housing and healthcare. While much of Paraguay’s Indigenous population lives in rural areas (9), similar barriers to TB diagnosis, treatment, and broader healthcare exist among Indigenous communities residing in urban areas.

In Paraguay, a previous study found that the relative risk of TB in prisons compared to the community was 70.3 (95% CI, 67.7-73.1) and an increasing percentage of country-wide TB case notifications occurred among incarcerated individuals: 7.1% in 2009 and 14.5% in 2018 (5). The result was a 250% increase in TB cases among the incarcerated population, from 2009 to 2018 (5).

The Indigenous population of Paraguay is another group with extremely high incidence of TB, estimated to be 335.3 per 100,000 in 2016 (7). This incidence translates to a relative risk of TB among Paraguay’s Indigenous population of 9.3, the highest relative risk associated with the Indigenous population among countries in the WHO Region of the Americas, three times higher than the next closest country (7). Additionally, over a quarter of Paraguay’s population falls below the national poverty line, potentially putting this group at heightened risk of TB (10).

Populations with heightened risk of TB in Paraguay have been identified in previous studies (5,7), but updated estimates of relative risk of TB among these populations is needed, as well as better understanding the spatial distribution of TB. Here, we analyzed TB notification data from the Paraguay National Program for Tuberculosis Control (NPTC) from 2018-2022 to identify populations with high incidence of TB that would benefit from increased access to TB prevention, diagnostics, and treatment. We identified populations and regions with high TB risk and evaluated socioeconomic predictors of treatment failure.

## METHODS

### Country setting

There are 7.1 million inhabitants of Paraguay, of which two percent are Indigenous (9). The country is divided by the Paraguay River into two main regions: the Eastern Region, which is home to 97% of the population, and the Western Region (also known as the Paraguayan Chaco), covering 61% of the national territory (14). The Western Region has a lower population density, mainly comprised of Indigenous communities, which account for 30.1% of the region’s population and represent 45.6% of the national Indigenous population. The country is divided into 17 departments. Asunción, the capital city, along with the Central Department, forms the country’s largest metropolitan area, including 37.6% of the national population (14). For this study, we refer to Asunción as the area including both the Department of Asunción and Central Department.

As recent estimates of the Indigenous population in each department were not available, we estimated department-level 2022 Indigenous populations using a linear model based on the official 2002 and 2012 census data to calculate department TB incidence among this population. All 17 prisons in Paraguay are located in the country’s Eastern Region. A 2022 prison census identified 17,006 incarcerated individuals, 94.8% of whom are men. This represented 0.22% of the national population in 2022 (15).

### NPTC Data Collection

Data on all TB patients were obtained from the NPTC database. As of 2018, the NPTC implemented the Tuberculosis Expert System (SEPNCT), which is a computer application component of the National Health Information System (SINAIS), intended to collect data related to suspected and confirmed cases of TB. The purpose of SEPNCT is to record and store information about patients, suspected cases, laboratory data, their treatments, monitoring, and contacts, for statistical, monitoring, surveillance, and notification purposes.

For this study, we focused on TB cases reported from January 2018 to December 2022.

The database is dynamic, that is, annual data from specific years uploaded to SEPNCT are continually updated and may change again in subsequent years. Socio-demographic variables were taken from the NPTC registry (sex, age, residence) of all notified TB patients, including those who belonged to TB risk groups under study, along with comorbidities, history of previous tuberculosis treatment, clinical-epidemiological variables, and treatment results. Analysis and data organization was completed in R (version 4.2.1) and R Studio (version 2023.03.1+446) using the *tidyverse* packages (11).

### Case Definitions

New TB cases were defined using the “history of previous treatment” variable. New patients were defined as having never received treatment for TB or having previously received treatment for less than one month. Previously treated patients were defined as having received treatment for one month or longer. Previously treated patients are further classified by the results of their most recent treatment cycle into: “Relapse,” “Treatment after failure,” “Treatment after loss to follow-up,” or “Other previously treated patients.” Relapse is defined as a previously treated patient, declared cured or treatment complete at the end of their last treatment cycle, and who is diagnosed with a subsequent episode of TB whether due to reactivation or reinfection (12,13). Treatment after failure refers to a patient with treatment failure at the end of their most recent treatment. Treatment after loss to follow-up refers to a patient declared lost to follow-up at the end of their most recent treatment, previously known as abandonment. Other previously treated patients are those previously treated, but with unknown or undocumented discharge results after the most recent treatment. We defined notified cases to include both “New patients” and “Relapsed patients.” This included 13,725 of the 15,025 total reported cases (91.3%).

The NPTC defines a person deprived of liberty as an individual who is reported to be incarcerated at the time of notification of the diagnosis of TB. Membership in an Indigenous population is self-reported to the NPTC. Indigenous and incarcerated population variables had no missing data, and only the responses “Yes” or “No.” The incarcerated population variable was cross checked with a “Penitentiary” binary variable (reporting if the patient was in a penitentiary) to ensure completeness.

Treatment success was determined using the “treatment result” variable. Patients reported as “cured” and “completed treatment” were considered to have successful treatment. Patients were considered cured if they completed treatment as recommended by national policy without evidence of failure and possessed three or more consecutive negative cultures with an interval of at least 30 days between them. Patients were recorded as having completed treatment if they had completed treatment as recommended by national policy without evidence of failure, but without evidence of three or more consecutive negative cultures at least 30 days apart.

### Mapping

Mapping was completed in R using the packages *sf* (16) and *ggplot2* (17). Paraguay department level shape files were obtained from the Paraguay – Subnational Administrative Boundaries dataset provided by the UN Office for the Coordination of Humanitarian Affairs “Humanitarian Data Exchange (18).” Map population data was determined by the “department of residence” category in the NPTC database, which reflects the home address that the patient provides. This is also adjusted in the information system, in case the patient moves, so they can receive their medication. This information is kept up to date in SEPNCT. All incident case observations had a listed department of residence. 5 of the 13,725 incident cases (0.04%) had a department of residence listed as “Foreign.” These five cases were excluded in population totals for mapping. General population data for mapping was obtained from the “Projection of the National Population” data provided by the Paraguay National Institute of Statistics (19).

### Statistical Analysis

Statistical analysis was carried out using R. T-tests and chi-square tests were performed using the *stats* package (20). Adjusted odds ratios were obtained from a linear model produced with the *glm* function from the stats package (20). Risk ratios for the incarcerated and Indigenous individuals were calculated with 2022 case notifications, and population data from the Paraguay National Institute of Statistics (19). We tested the effect on treatment outcome of clinical and sociodemographic variables in a multivariable logistic regression model that included age, gender, education level, year of case diagnosis, incarceration status, enrollment in directly observed therapy (DOT), self-reported Indigenous population status, diabetes, alcohol use disorder, mental disorder, HIV status, possession of a GeneXpert test result, and an interaction term between incarceration status and DOT. A total of 12% of notified cases had a treatment result that was either missing or in progress. These notifications were excluded in the linear model. An additional 43 cases were reported to be positive for rifampin resistance and were also excluded from treatment outcome analysis.

### Ethics Statement

This study was approved by the IRB at the University of Utah (IRB_00167535) and by the Instituto de Medicina Tropical, Asunción, Paraguay.

## RESULTS

From 2018-2022, a total of 13,725 incident TB cases were reported by the National Program for Tuberculosis Control (NPTC) in Paraguay. Countrywide case notification rates remained constant over this five-year period at approximately 46 cases per 100,000 (χ^2^ test, p=0.78). Of the 13,725 total reported TB cases, 1,743 (12.7%) were among the Indigenous population and 2,331 (17%) were among the incarcerated population.

We found significant variation in case notification across individual departments (X^2^ test, p<0.001) and variation in trends over time. Over the 5-year study period, the highest increase in notification was in Boquerón, where case notifications rose from 118.2 per 100,000 in 2018 to 157.3 per 100,000 in 2022 (Fig. 1). Other departments that experienced notable increases were Concepción (an increase of 94.5%), Amambay (63.0%), and Cordillera (63.1%). The greatest decrease in notifications was in Alto Paraguay, where case notifications decreased from 96.9 per 100,000 in 2018 to 73.9 per 100,000 in 2022. Boquerón and Alto Paraguay are both departments in the Chaco with a high proportion of Indigenous residents, 43.4% and 25.1%, respectively. In total, 45% of TB cases (787 out of 1,743) among the Indigenous population were located in the Chaco Region (Fig. 2). Among the TB cases in the Chaco, approximately 75% (787 out of 1,048) were among individuals identifying as Indigenous.

**Figure 1:**
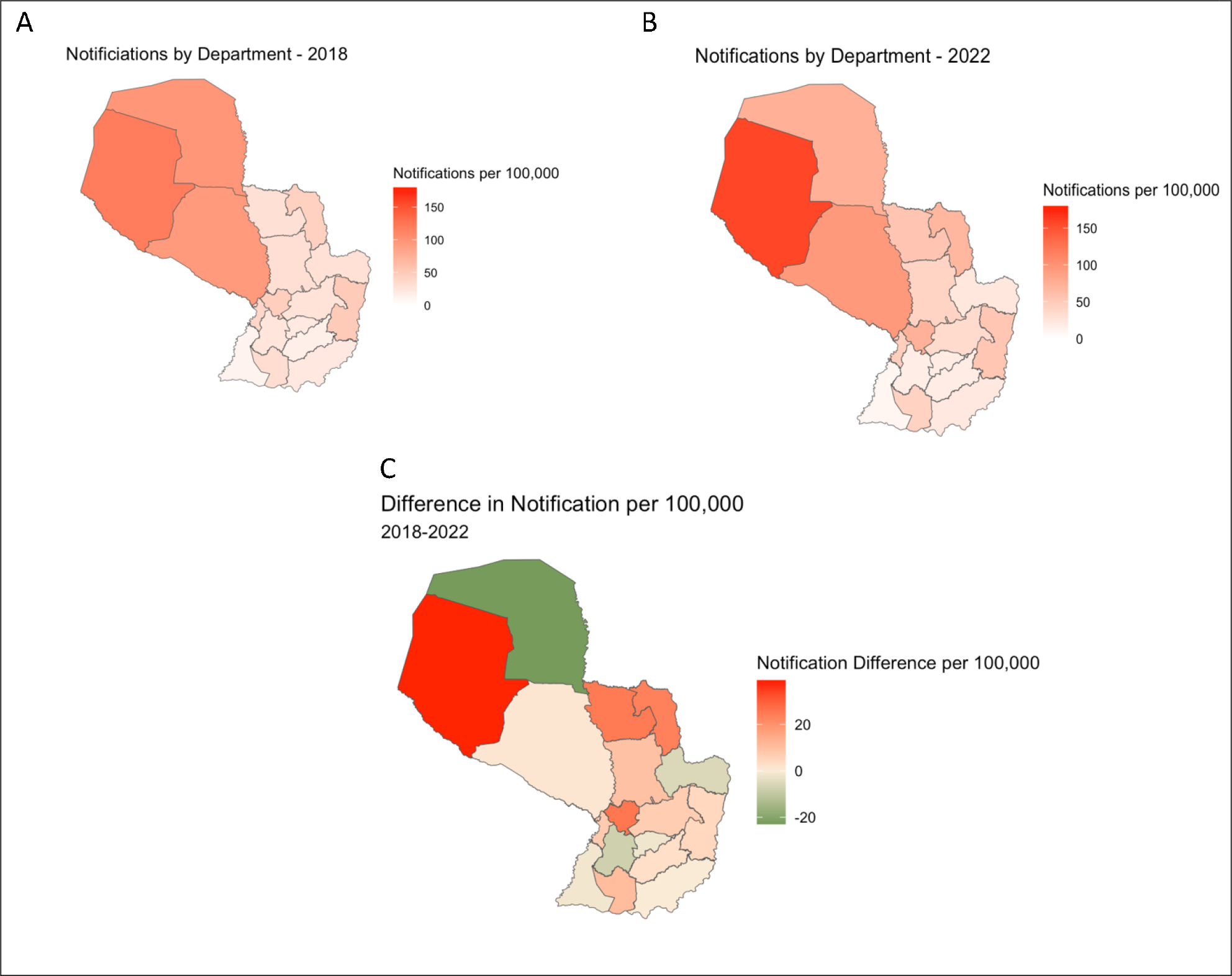
Tuberculosis Notifications in Paraguay 2018-2022 TB notifications per 100,000 in Paraguay by department in (A) 2018 and (B) 2022. (C) Difference from 2018 to 2022 in tuberculosis notification per 100,000 by department.

**Figure 2:**
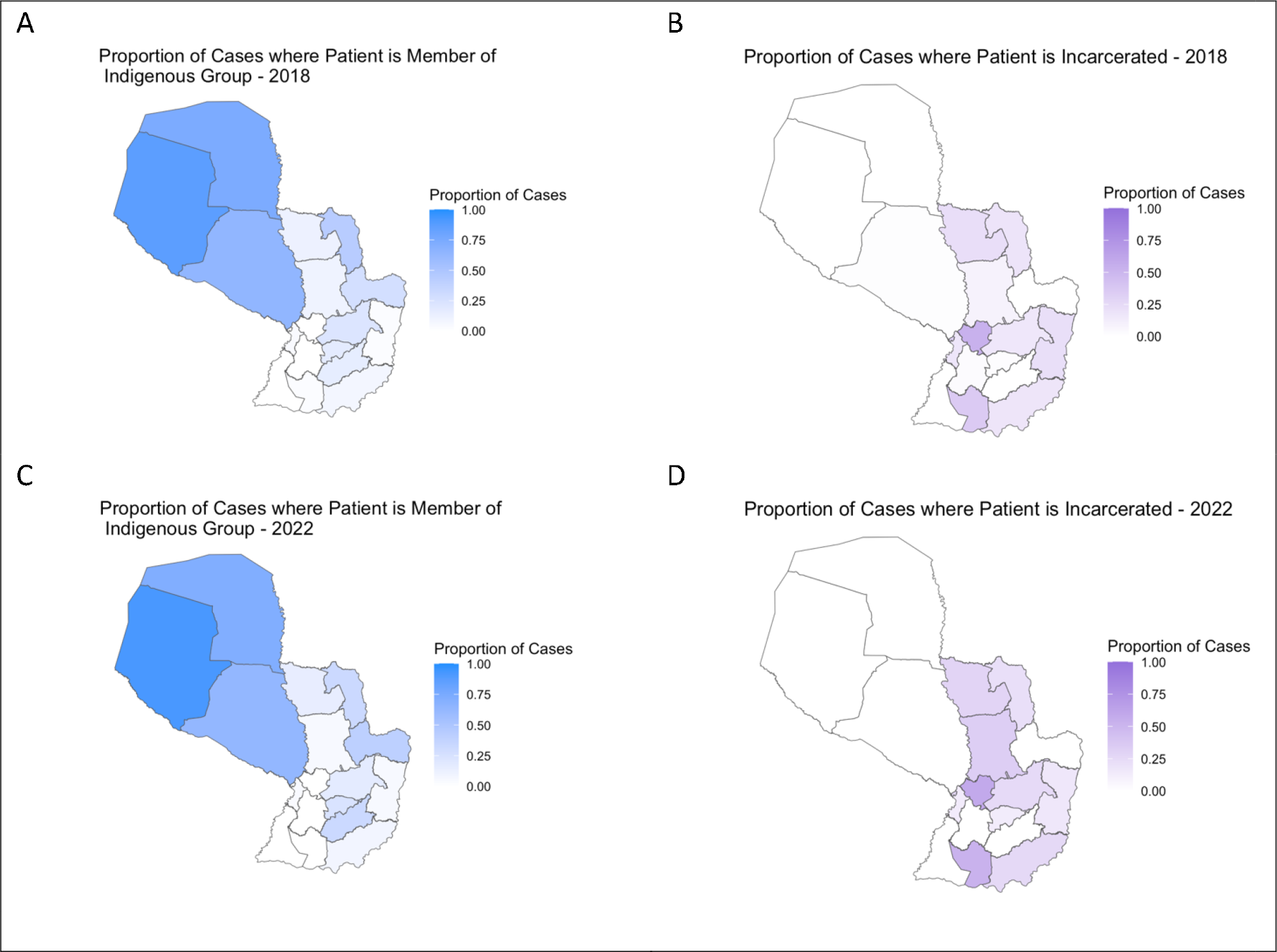
Proportion of Tuberculosis Cases in Paraguay Among Indigenous and Incarcerated Populations Proportion of reported tuberculosis cases by department that indicated being a member of an Indigenous group in (A) 2018, (C) 2022, and that are incarcerated in (B) 2018, (D) 2022.

We observed significant variation in TB notifications in the Indigenous population across space. Asunción has the highest notification rate among the Indigenous population of 1127.4 cases per 100,000 in 2022 (Fig. 3). Other departments also had high notification rates: 793.7 cases per 100,000 in Guairá and 493.8 cases per 100,000 in Itapúa in 2022. In 2022 Paraguay’s Indigenous populations had a relative risk of TB notification 6.4 times higher than the non-Indigenous population.

**Figure 3:**
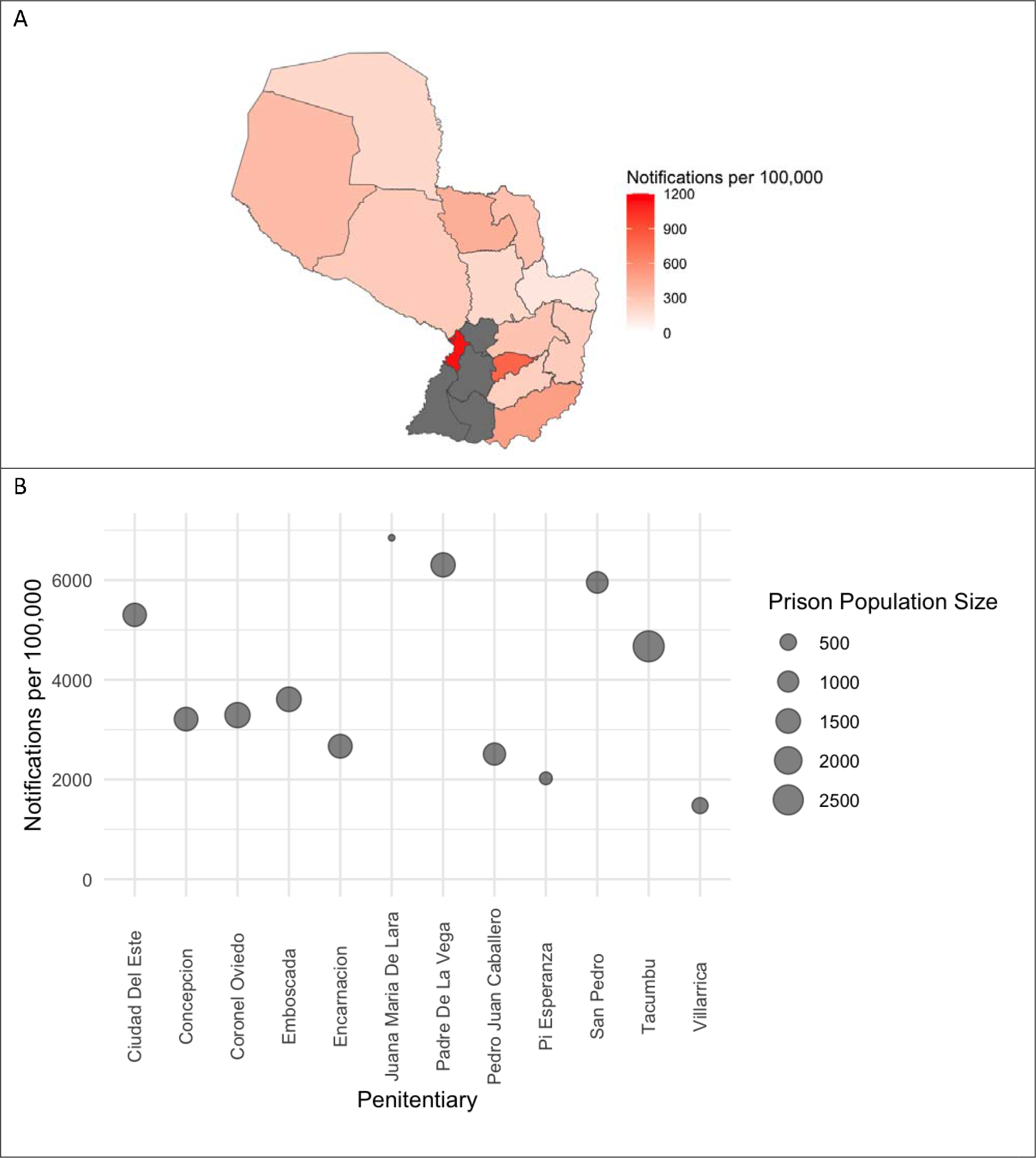
Tuberculosis Among Paraguay’s Vulnerable Populations in 2022 (A) 2022 TB notifications per 100,000 among the Indigenous population (departments in grey had no reported Indigenous residents), and (B) 2022 TB notifications and population size of major prison facilities^1^.

Among the Indigenous population, 54.4% (948 out of 1743) of reported cases occurred in men. The proportion of individuals with TB who were HIV co-infected was lower among the Indigenous population than the non-Indigenous population (1.7% vs 7.6%, p<0.001). Individuals diagnosed with TB who are Indigenous had a lower median age than their non-Indigenous counterparts (32 years vs. 35 years, p<0.001).

2,331 of the 13,725 reported TB cases (17%) were among the incarcerated population. The departments with the highest proportion of cases among the incarcerated population were Cordillera (55.4%, 456 of 823) and Misiones (44.2%, 91 of 206). TB cases among the incarcerated population occur in the Eastern Region, where all Paraguay’s prisons are located (Fig. 2). The majority of cases notified among the incarcerated population occurred in men, 98.9%, compared with 67.1% in the general population (X^2^ test, p<0.001). The proportion of individuals with TB who were HIV co-infected was lower among the incarcerated population than the general population (3.5% vs 7.6%, p<0.001). The median age of incarcerated individuals with TB was lower (28 vs. 35, p<0.001) and people who are incarcerated were more likely to be treated under DOT (88.7% vs 53.6%, p<0.001) than non-incarcerated individuals with TB. In 2022, Paraguay’s incarcerated population had a relative risk of TB notification 87 times higher than the non-incarcerated population.

Rapid molecular tests, including GeneXpert, are the recommended diagnostic test for TB diagnosis (21). From 2018-2022, GeneXpert usage for diagnosing incident cases of TB increased from 23% in 2018 to 72% in 2022 (t-test; p<0.001), and a pattern of increase was observed across subpopulations (Fig. 4). From 2018-2022, the proportion of TB cases diagnosed with GeneXpert among the Indigenous population was slightly lower than that diagnosed using GeneXpert among the non-incarcerated, non-Indigenous population (35.4% vs. 37.0%, p<0.001). TB cases among the incarcerated population were diagnosed using GeneXpert at a higher rate than the non-incarcerated, non-Indigenous population (55.3% vs. 37.0%, p<0.001).

**Figure 4:**
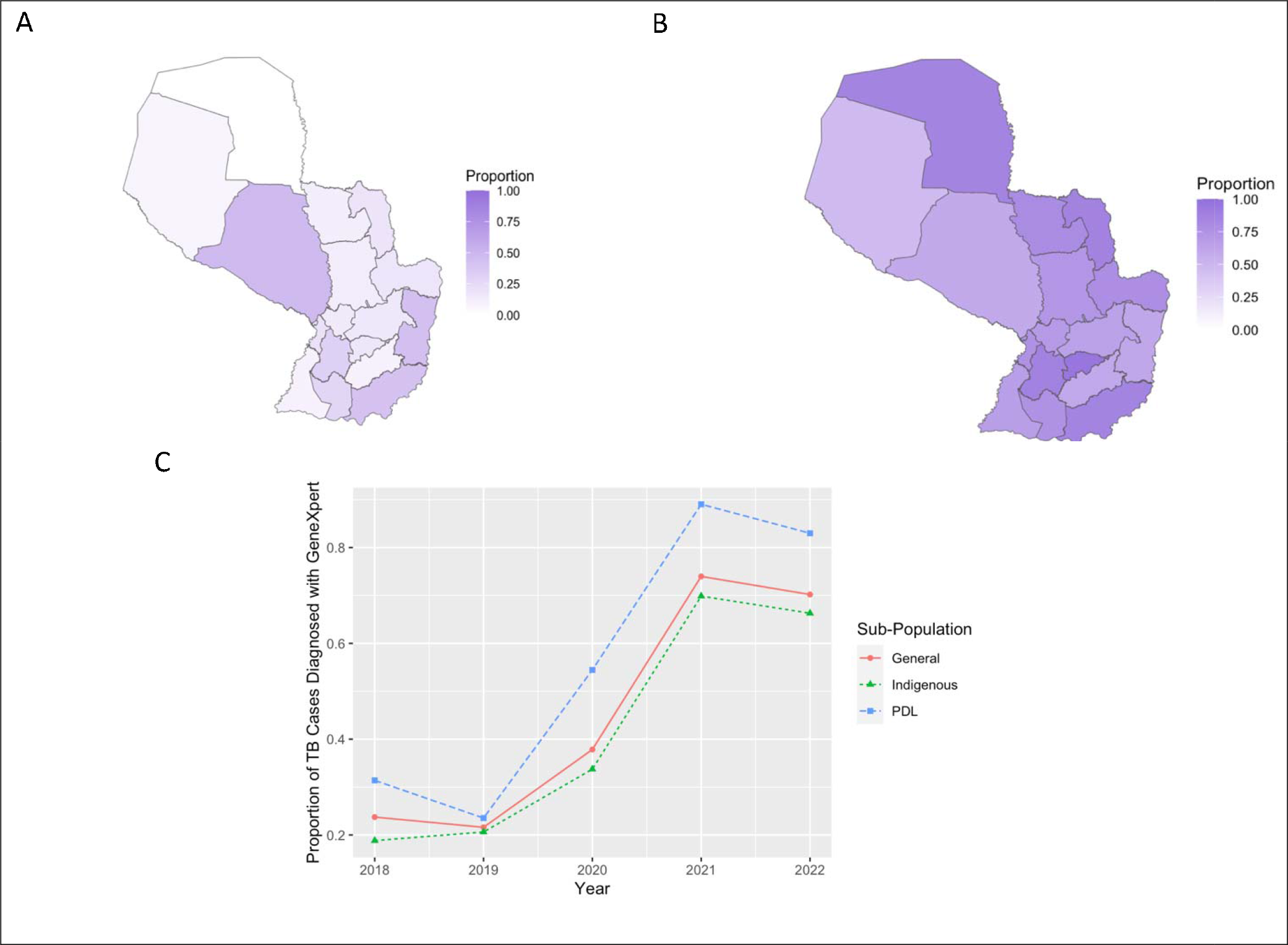
GeneXpert Testing Usage in Paraguay, 2018-2022 Proportion of tuberculosis cases in Paraguay with a GeneXpert result by department in 2018 (A), and 2022 (B). (C) Proportion of tuberculosis cases in Paraguay with a GeneXpert result divided by subpopulation from 2018 to 2022.

Of the 13,725 notified TB cases recorded by the Paraguay NPTC from 2018-2022, 8,238 (60%) had successful treatment, 2,240 (16%) were lost to follow up, 1,451 (11%) died, 85 (0.6%) reported treatment failure, 1,293 (9%) reported treatment still in progress, and 418 (3%) were missing data.

In a multiple linear regression model (Table 2), DOT (Directly Observed Therapy) (aOR: 1.66; 95% CI: 1.51, 1.84), diabetes (aOR: 1.39; 95% CI: 1.17-1.66), being a member of the Indigenous population (aOR: 1.41; 95% CI: 1.21-1.66), and Tertiary/University level education (aOR: 3.68; 95% CI: 2.85-4.78) were positively associated with treatment success for drug-susceptible TB. While incarceration alone was not significantly associated with positive or negative treatment success (aOR: 1.15; 95% CI: 0.81–1.64), an interaction term between DOT and incarceration was positively associated with successful treatment (aOR: 2.51; 95% CI: 1.69- 3.70). Factors negatively associated with treatment success included co-infection with HIV (aOR: 0.20; 95% CI: 0.17-0.23), Alcohol Use Disorder (aOR: 0.70; 95% CI: 0.59-0.84), and age over 65 (aOR: 0.42; 95% CI: 0.34-0.51).

**Table 1.**
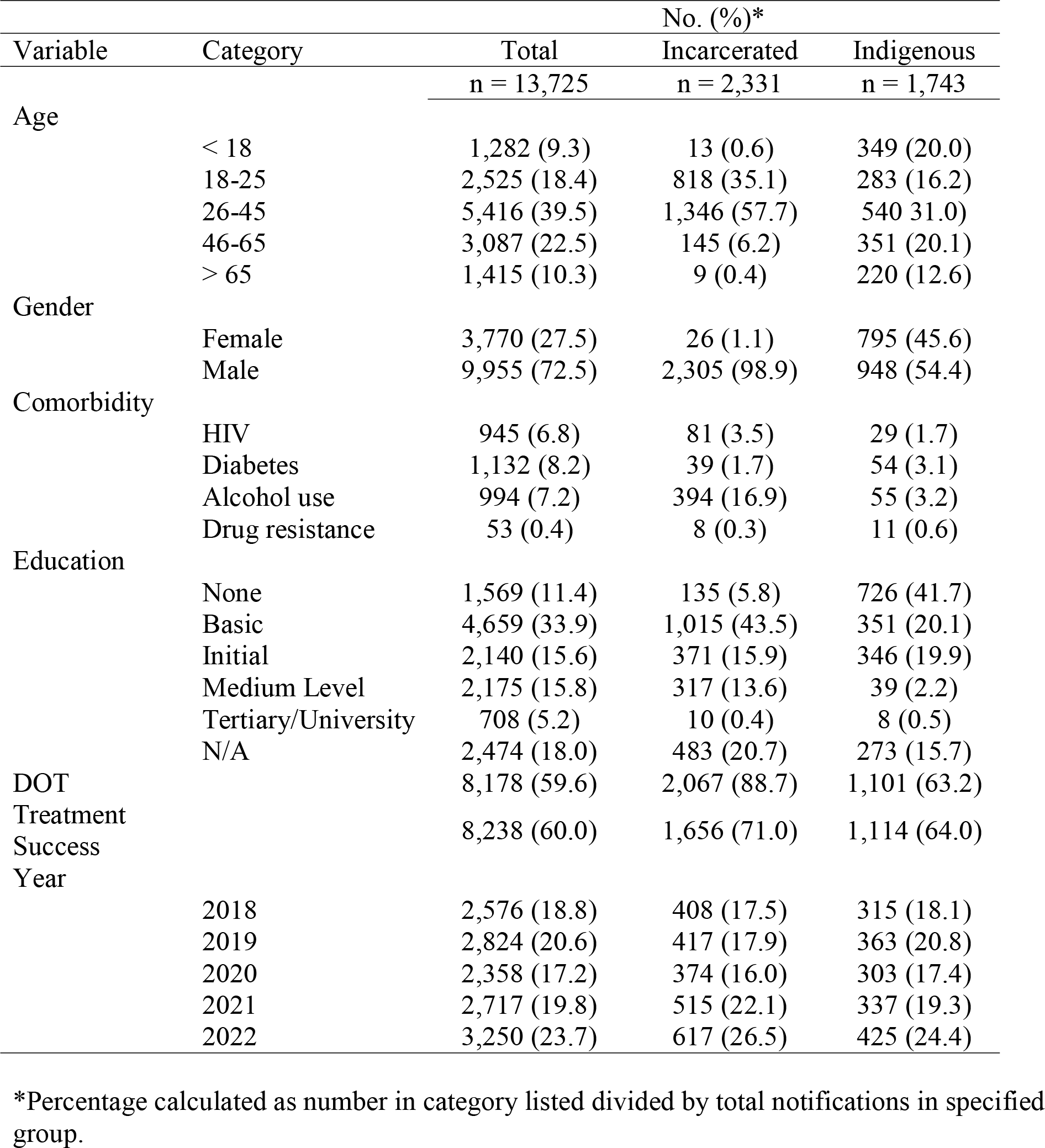
Sociodemographic characteristics for new TB case notifications 2018-2022, by incarcerated and Indigenous status.

**Table 2.** Adjusted odds ratios for treatment success among all newly diagnosed Tuberculosis cases using multiple linear regression.

| Variable | Category | Adjusted Odds Ratio (95% CI) |
| --- | --- | --- |
| Directly Observed Therapy |  | 1.66 (1.51 – 1.84) |
| Incarceration |  | 1.15 (0.81 – 1.64) |
|  | Incarceration*DOT<sup>2</sup> | 2.51 (1.69 – 3.70) |
| Comorbidities |  |  |
|  | HIV Positive | 0.20 (0.17 – 0.23) |
|  | Diabetes | 1.39 (1.17 – 1.66) |
|  | Alcohol Use Disorder | 0.70 (0.59 – 0.84) |
| Member of Indigenous population |  | 1.41 (1.21 – 1.66) |
| Age |  |  |
|  | < 18 | Referent |
|  | 18-25 | 0.62 (0.50 – 0.75) |
|  | 26-45 | 0.66 (0.55 – 0.79) |
|  | 46-65 | 0.63 (0.52 – 0.76) |
|  | > 65 | 0.42 (0.34 – 0.51) |
| Education |  |  |
|  | None | Referent |
|  | Basic | 1.65 (1.42 – 1.92) |
|  | Initial | 1.87 (1.58 – 2.21) |
|  | Medium Level | 2.05 (1.72 – 2.44) |
|  | Tertiary/University | 3.68 (2.85 – 4.78) |

<sup>2</sup> Incarceration\*DOT indicates the use of an interaction term

## DISCUSSION

Despite advances in diagnosis and treatment, TB remains a significant public health threat in Paraguay, with a disproportionate impact among populations marginalized from healthcare, including Indigenous people and incarcerated individuals. In this retrospective study conducted between 2018 and 2022, based on NPTC data, we found that TB case notifications are concentrated spatially and within specific subpopulations.

Paraguay’s Indigenous population continues to have an exorbitantly high relative risk of TB. Our findings confirm that the Chaco Region, the region home to the majority of Paraguay’s Indigenous population, has the highest incidence of TB in the country. In 2017, members of the Pan American Health Association developed a shared set of guidelines to improve Indigenous health, including reducing the burden of TB (7). Since 2019, the NPTC has expanded access to diagnostics and treatment and treatment support programs for Indigenous populations, which may be responsible for the improvements in treatment success we observe here.

The TB epidemic in Paraguay is also increasingly concentrated in prisons, where the incidence of TB among the incarcerated population remains a major health justice issue. Most TB cases among the incarcerated population are located in the eastern part of the country, including the departments of Cordillera and Misiones. We found that while the use of DOT increases odds of treatment success for everyone, it is particularly important for the incarcerated population. This is consistent with a study in Brazil, where DOT access was associated with greater improvements in treatment outcomes among incarcerated individuals compared to the non-incarcerated population (22). Recently, the NPTC has implemented TB programs in penitentiary centers that aim to provide universal DOT. Our findings emphasize the broad importance of access to DOT, and the need for its continued expansion within populations with the highest incidence of TB.

Paraguay was one of the first countries in the WHO Americas Region to adopt molecular rapid tests for initial TB diagnosis. As of October 2020, GeneXpert testing replaced sputum microscopy as the initial TB diagnostic. Previously, GeneXpert testing was applied as part of targeted recommendations for groups with high incidence, including incarcerated individuals. We found significant increases in GeneXpert use country-wide, including in incarcerated and Indigenous populations. Historically, access to TB diagnostics was limited in the Chaco, leading to what was likely a large burden of undiagnosed TB. We found that all case notifications among the Indigenous population in Alto Paraguay, one of the three departments in the Chaco, possessed a GeneXpert testing result.

We expect that the ongoing expansion of GeneXpert testing will continue to improve case detection rates, critical for both improving TB treatment outcomes and reducing transmission.

We found that rates of access to GeneXpert testing availability among the Indigenous population lag behind the non-Indigenous population, highlighting the need to prioritize the expansion of diagnostics to this population in both the Chaco Region and the urban Indigenous population in the Central Department, which has the highest incidence rate among Indigenous peoples.

Our analysis was made possible by the detailed data provided by the NPTC. Comprehensive data that was geographically tracked and connected to sociodemographic indicators provides valuable insight into TB trends over time. Additionally, it allows for spatially informed analysis that guides effective interventions to areas of high need.

Our study has several limitations. Our analysis of incidence rates among the Indigenous population relied on Indigenous population data. At the time of analysis, the last official census data including Indigenous population data was from 2012. To calculate 2022 incidence rates, we used linear regression to estimate the 2022 Indigenous population data using the official 2002 and 2012 census data.

Further, we were unable to examine the spatial distribution of TB cases at a resolution higher than department-level because of missing information in the TB registry. However, we were still able to identify meaningful spatial patterns. Additionally, the TB registry lacked information about prior incarceration. Incarceration history is a major risk factor for TB and it is possible that many cases that were notified among non-incarcerated individuals occurred among individuals with recent incarceration. Lastly, working with reported case data presents the challenge that we cannot estimate unreported cases. Unreported cases may occur disproportionately among vulnerable populations, leading us to underestimate incidence rates.

TB remains a major public health problem in Paraguay, with a burden that is concentrated among vulnerable populations. Our study highlights the benefit of expanding molecular TB testing across the country and highlights areas and populations that could benefit from continued expansion of molecular testing. We also found that DOT treatment continues to be a critical component of treatment regimes, particularly for incarcerated populations. Together, these findings highlight the urgency in supporting public health programs to improve the diagnosis, treatment, and prevention of TB among groups historically marginalized from health and social services to reduce the exceedingly high burden of TB.

## Data Availability

All data produced in the present study are available upon reasonable request to the authors.

## Footnotes

1 San Juan Bautista was excluded due to unavailable population data

